# Population-scale Longitudinal Mapping of COVID-19 Symptoms, Behavior, and Testing Identifies Contributors to Continued Disease Spread in the United States

**DOI:** 10.1101/2020.06.09.20126813

**Authors:** William E. Allen, Han Altae-Tran, James Briggs, Xin Jin, Glen McGee, Andy Shi, Rumya Raghavan, Mireille Kamariza, Nicole Nova, Albert Pereta, Chris Danford, Amine Kamel, Patrik Gothe, Evrhet Milam, Jean Aurambault, Thorben Primke, Weijie Li, Josh Inkenbrandt, Tuan Huynh, Evan Chen, Christina Lee, Michael Croatto, Helen Bentley, Wendy Lu, Robert Murray, Mark Travassos, Brent A. Coull, John Openshaw, Casey S. Greene, Ophir Shalem, Gary King, Ryan Probasco, David R. Cheng, Ben Silbermann, Feng Zhang, Xihong Lin

**Author notes:** These authors contributed equally (William E. Allen, Han Altae-Tran, James Briggs, Xin Jin, Glen McGee, Andy Shi).

## Abstract

Despite social distancing and shelter-in-place policies, COVID-19 continues to spread in the United States. A lack of timely information about factors influencing COVID-19 spread and testing has hampered agile responses to the pandemic. We developed How We Feel, an extensible web and mobile application that aggregates self-reported survey responses, to fill gaps in the collection of COVID-19-related data. How We Feel collects longitudinal and geographically localized information on users’ health, behavior, and demographics. Here we report results from over 500,000 users in the United States from April 2, 2020 to May 12, 2020. We show that self-reported surveys can be used to build predictive models of COVID-19 test results, which may aid in identification of likely COVID-19 positive individuals. We find evidence among our users for asymptomatic or presymptomatic presentation, as well as for household and community exposure, occupation, and demographics being strong risk factors for COVID-19. We further reveal factors for which users have been SARS-CoV-2 PCR tested, as well as the temporal dynamics of self-reported symptoms and self-isolation behavior in positive and negative users. These results highlight the utility of collecting a diverse set of symptomatic, demographic, and behavioral self-reported data to fight the COVID-19 pandemic.

## Main Text

The rapid global spread of severe acute respiratory syndrome coronavirus 2 (SARS-CoV-2), the novel virus causing coronavirus disease 2019 (COVID-19)^1–3^, has created an unprecedented public health emergency. In the United States, efforts to slow the spread of disease have included, to varying extents, social distancing, home-quarantine and treating infected patients, mandatory facial covering, closure of schools and non-essential businesses, and testing-trace-isolate measures^4,5^. The COVID-19 pandemic and ensuing response has produced a concurrent economic crisis of a scale not seen for nearly a century^6^, exacerbating the effect of the pandemic on different socioeconomic groups and producing adverse health outcomes beyond COVID-19. As a result, there is currently intense pressure to safely wind down these measures. Yet, in spite of widespread lockdowns and social distancing throughout the US, many states continue to exhibit steady increases in the number of cases^7^. In order to understand where and why the disease continues to spread, there is a pressing need for real-time individual-level data on COVID-19 infections and tests, as well as on the behavior, exposure, and demographics of individuals at the population scale with granular location information. These data will allow medical professionals, public health officials, and policy makers to understand the effects of the pandemic on society, tailor intervention measures, efficiently allocate testing resources, and address disparities.

One approach to collecting this type of data on a population scale is to use web- and mobile-phone based surveys that enable large-scale collection of self-reported data. Previous studies, such as FluNearYou, have demonstrated the potential for using online surveys for disease surveillance^8^. Since the start of the COVID-19 pandemic, several different applications have been launched throughout the world to collect COVID-19 symptoms, testing, and contact-tracing information^9^. Studies in the UK and Israel have reported large cohorts of users and demonstrated some ability to detect and predict the spread of disease^10–12^. Existing tools, however, focus primarily on COVID-19 symptoms. There is a strong need to investigate exposure, demographic and behavioral factors that affect the chain of transmission, understand the factors for who have been tested, and study the degree of presence of asymptomatic, presymptomatic, mildly symptomatic cases^13^.

To overcome these limitations, we developed How We Feel (HWF, http://www.howwefeel.org) (Fig. 1a-d), a web and mobile-phone application for collecting de-identified self-reported COVID-19-related data. Rather than targeting suspected COVID-19 patients or existing study cohorts, HWF aims to collect data from users representing the population at large. Users are asked to share information on demographics (gender, age, race/ethnicity, household structure, ZIP code), COVID-19 exposure, and pre-existing medical conditions. They then self-report daily how they feel (well or not well), any symptoms they may be experiencing, test results, behavior (e.g., use of face coverings), and sentiment (e.g., feeling safe to go to work) (Fig. 1c, Extended Data Fig. 1). To protect privacy, users are not identifiable beyond a randomly-generated number that links repeated logins on the same device. A key feature of the app is the ability to rapidly release revised versions of the survey as the pandemic evolves. In the first month of operation, we released three iterations of the survey with increasingly expanded sets of questions (Fig. 1b).

**Figure 1:**
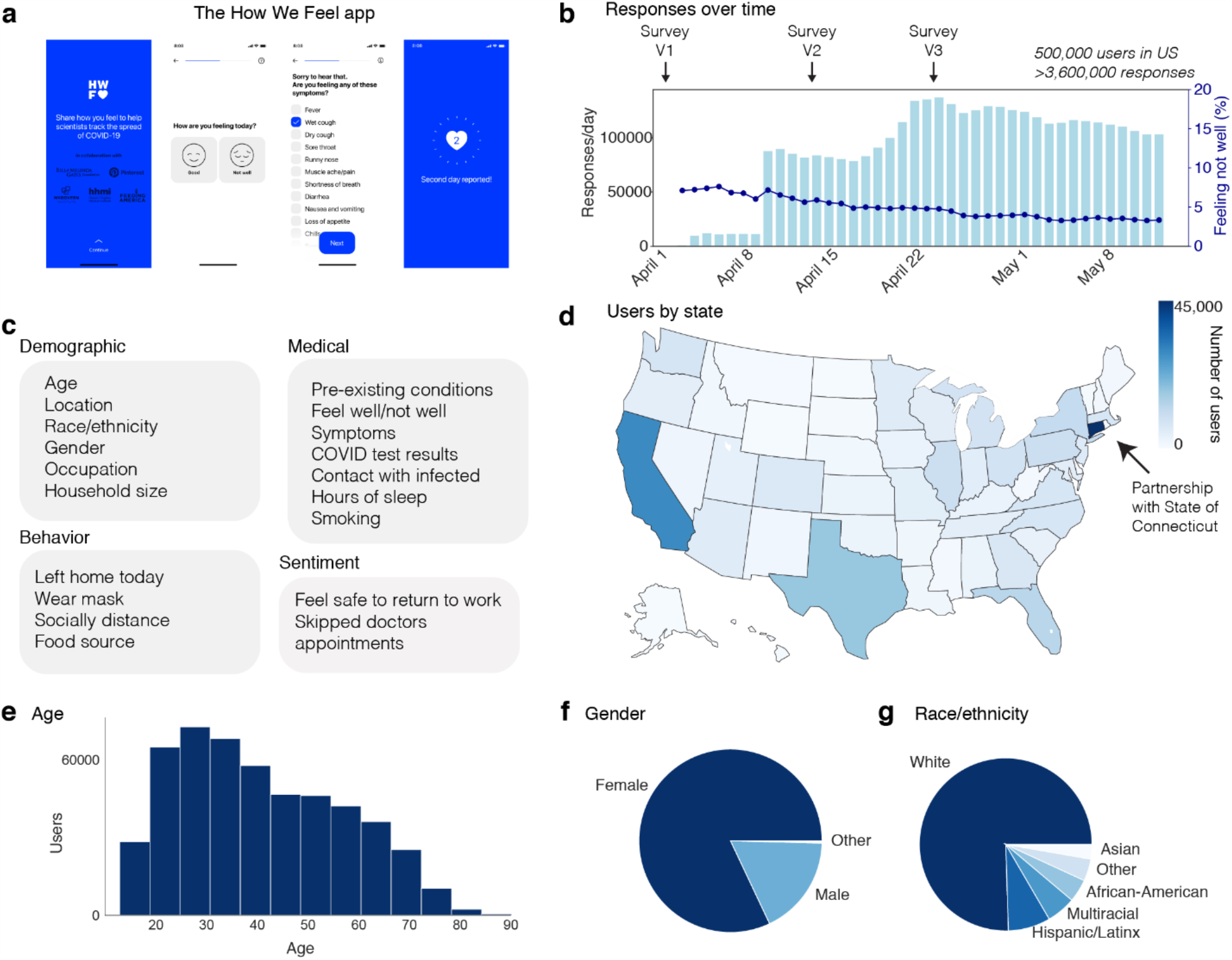
The How We Feel Application and User Base. **a**, The How We Feel (HWF) app: longitudinal tracking of self-reported COVID-19-related data. **b**, Responses over time, as well as percentage of users reporting feeling unwell, with releases of major updates to survey indicated. **c**, Information collected by the HWF app. **d**, Users by state across the United States. **e**, Age distribution of users. Note: users had to be older than 18 to use the app. **f**, Distribution of self-reported sex. **g**, Distribution of self-reported race or ethnicity. Users were allowed to report multiple races. “Multiracial” = the user indicated more than one category. “Other” includes American Indian/Alaskan Native and Hawaiian/Pacific Islander, as well as users who selected “Other”.

The app was launched on April 2, 2020 in the United States. As of May 12, 2020, the app had 502,731 users in the United States, with 3,661,716 total responses (Fig. 1b) (Extended Data Table 1). 74% of users responded on multiple days, with an average of 7 responses per user (Extended Data Fig. 2). Each day, ∼5% of users who accessed the app reported feeling unwell (Fig. 1b). The user base was distributed across all 50 states and several US territories, with the largest numbers of users in more populous states such as California, Texas, Florida, and New York (Fig. 1d). Connecticut had the largest number of users per state, as the result of a partnership with the Connecticut state government (Fig. 1d). Users were required to be 18 years of age or older and were 42 years old on average (mean: 42.0; SD: 16.3), including 18.4% in the bracket of 60+, which has experienced the highest mortality rate from COVID-19 (Fig. 1e)^14,15^. Users were primarily female (82.7%) (Fig. 1f) and white (75.5%, excluding 20.3% with missing data) (Fig. 1g).

**Figure 2:**
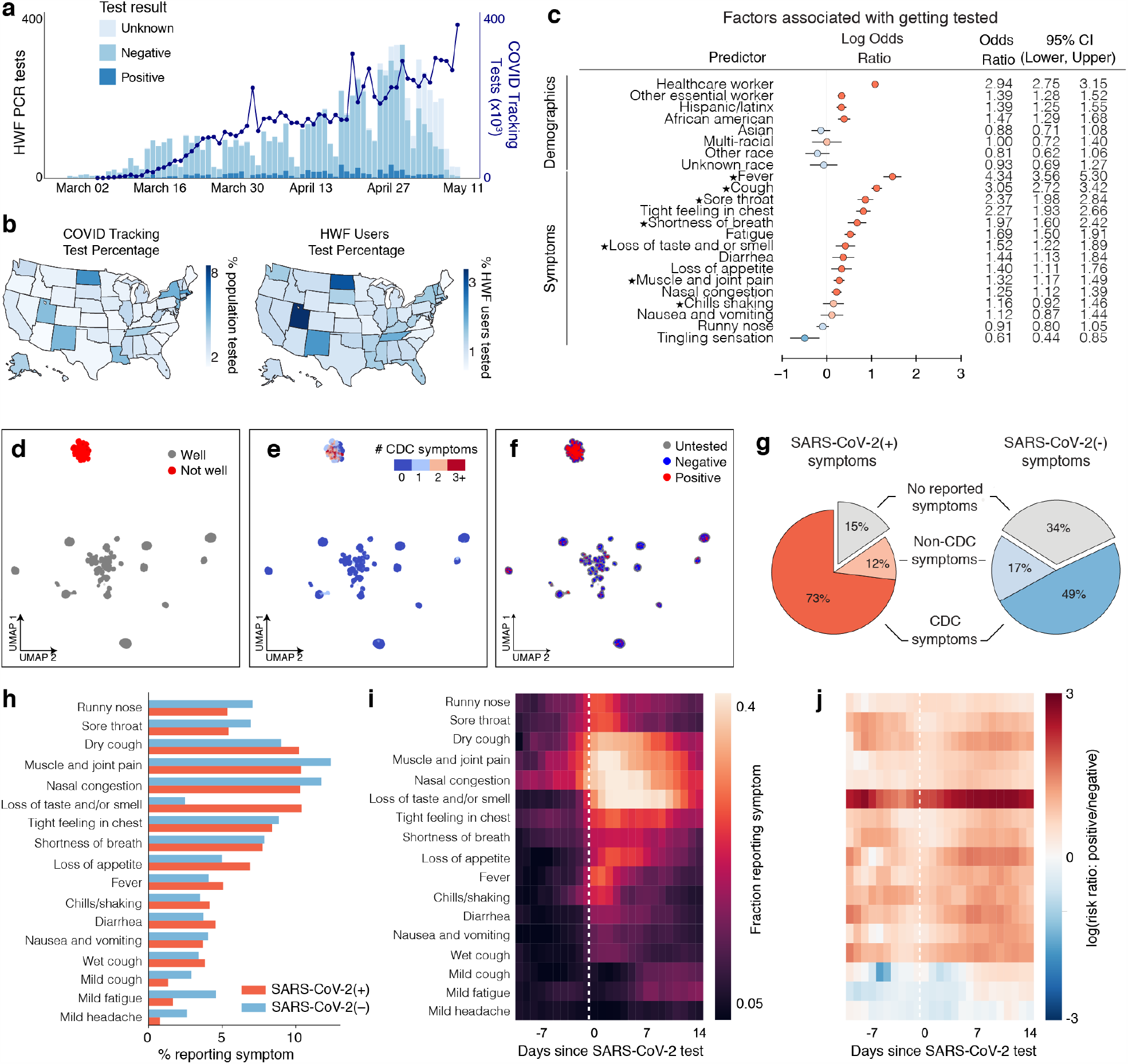
SARS-CoV-2 PCR Testing and Symptoms. **a**, Stacked bar plot of user-reported test results over time, overlaid with official number of tests across US based on COVID Tracking Project data. **b**, Left: Map of per-capita test rates across the United States. Right: Map of COVID-19 tests per number of users by state. **c**, Associations of professions and symptoms with receiving a SARS-CoV-2 PCR test, adjusted for demographics and other covariates (Methods). Common symptoms listed by the CDC are starred. **d-f**, UMAP visualization of 667,651 multivariate symptom responses among HWF users that reported at least one symptom. Coloring indicates: **d**, responses according to users feeling well; **e**, the reported number of COVID-19 symptoms listed by the CDC; and **f**, the COVID-19 test result among tested users. **g**, Proportion of positive COVID-19 patients (red) and negative COVID-19 patients (blue) experiencing either CDC-common symptoms (dark), only non-CDC symptoms (light), or no symptoms (grey) on the day of their test. **h**, Histogram of reported symptoms among COVID-19 tested users. **i**, Longitudinal self-reported symptoms from users that tested positive for COVID-19. Dates are centered on the self-reported test-date. **j**, Ratio of symptoms comparing users that test positive versus test negative for COVID-19.

A major ongoing problem in the US is the overall lack of testing across the country^16^ and disparities in test accessibility, infection rates, and mortality rates in different regions and communities^17,18^. In the absence of population-scale testing, it will be critical during a reopening to allocate limited testing resources to the groups or individuals most likely to be infected in order to track the spread of disease and break the chain of infection. We therefore first examined who in our userbase is currently receiving testing. We analyzed 4,759 users who took the Version 3 (V3) survey and who were PCR tested for SARS-CoV-2 (out of 272,392 total users) (Fig. 2a, Extended Data Fig. 3a). Of these, 8.8% were PCR positive. The number of tests reported by test date displays a similar trend to the estimated number of tests across the US, suggesting that our sampling captures the increase in test availability (Fig. 2a). The number of PCR tests per HWF user is highly correlated with external estimates of per-capita tests by state (Fig. 2b, Extended Data Fig. 3b, Pearson correlation 0.77)^19^.

**Figure 3:**
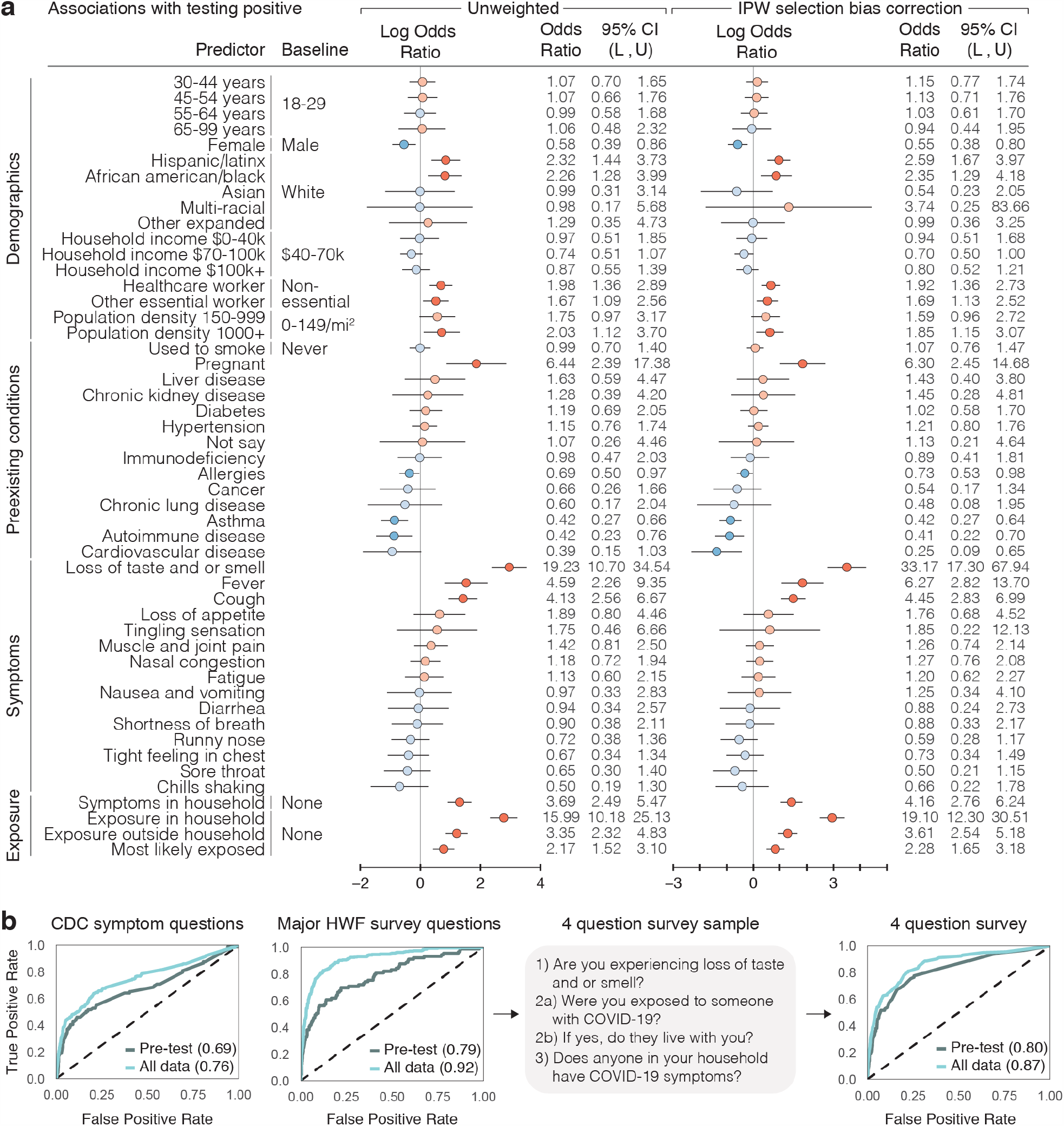
SARS-CoV-2 PCR Test Result Associations and Predictions. **a**, Factors associated with respondents receiving and reporting a positive test result, as determined through logistic regression. Left: results from unweighted model. Right: results from model incorporating propensity scores via inverse probability weights (IPW). Reference categories are indicated where relevant, and when not indicated, the reference is not having that specific feature. Log odds ratios and their confidence intervals are plotted, with red indicating positive association and blue indicating negative association. Darker colors indicate confidence intervals that do not cover 0. Population density and neighborhood household income were imputed from user demographics. L = lower bound, U = upper bound of 95% confidence intervals. **b**, Prediction of positive test results using ±2 weeks of data from test date, using 5-fold cross validation, shown as receiver operating characteristic (ROC) curves. The XGBoost model was trained on different subsets of questions: CDC Symptom Questions = using just the subset of COVID-19 symptoms listed by the CDC. All Survey Questions = using the entire survey. 4 Question survey = using a reduced set of 4 questions that were found to be highly predictive. Numerical values are AUC = area under the ROC curve.

We first examined via logistic regression which factors either collected in the survey or inferred from US Census data by user ZIP code were associated with receiving a SARS-CoV-2 PCR test, regardless of test result. As expected, we observed that a higher fraction of tested users from states with higher per-capita test numbers, according to the COVID Tracking Project^19^ (Extended Data Fig. 3b). Healthcare workers (OR: 2.94; 95% CI: [2.75, 3.15]) and other essential workers (OR: 1.39; 95% CI: [1.28, 1.52]) were more likely to have received a PCR test compared to users who did not report those professions (Fig. 2c). Users who reported experiencing fever, cough, or loss of taste/smell (among other symptoms) had higher odds of being tested compared to users who never reported these symptoms (Fig. 2c). The majority of these symptoms are listed as common for COVID-19 cases by the Centers for Disease Control and Prevention (CDC) (Fig. 2c, starred)^20^. A less common symptom, reporting a tight feeling in one’s chest, was also associated with receiving a PCR-based test (OR: 2.27, 95% CI: [1.93, 2.66]). These results suggest that the most commonly reported symptoms are being used as screening criteria for determining who receives a test, potentially missing individuals with less common symptom presentations. This group could include those who are at high risk for infection but do not meet the testing eligibility criteria.

To obtain a global view of self-reported symptom patterns, we applied an unsupervised manifold learning algorithm to visualize how symptoms were correlated across users (Methods). As expected, we found that symptom presentation separated broadly by feeling well versus feeling unwell (Fig. 2d, Extended Data Fig. 4). Users who felt unwell were concentrated in a single cluster indicating similar overall symptom profiles, which was characterized by high proportions of common COVID-19 symptoms as defined by the CDC^20^ (Fig. 2e), and contained the vast majority of responses from users with both positive (+) and negative (–) SARS-CoV-2 PCR tests (Fig. 2f). Thus COVID-19 symptoms tend to overlap with symptoms for other diseases and do not necessarily predict positive test results.

**Figure 4:**
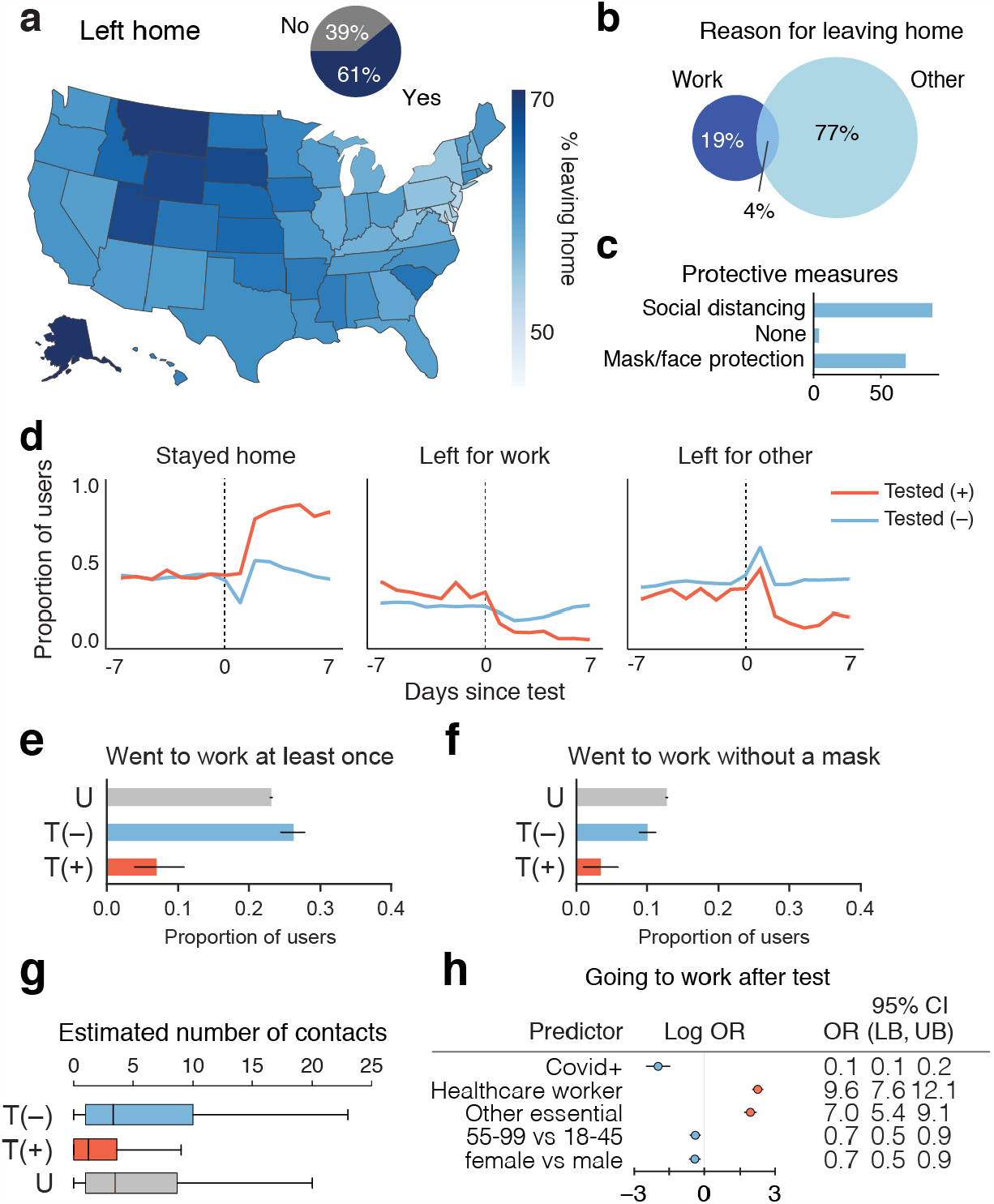
Behavioral Factors Potentially Contributing to COVID-19 Spread. **a**, Proportion of responses indicated users leaving home across US (map) or overall (inset pie chart). **b**, Percentage of responses of users reporting work or other reason for leaving home. **c**, Reported protective measures taken per response taken by users upon leaving home. **d**, Time course of proportion of SARS-CoV-2 PCR tested positive (+) or negative (–) users staying home, leaving for work, and leaving for other reasons. **e-f**, Proportion SARS-CoV-2 PCR tested (+) or (–), or untested (U), going to work (**e)**, going to work without a mask (**f**) in the 2-7 days post test for T = tested, or 3 weeks since last check in for U = untested. Healthcare workers and other essential workers are compared to non-essential workers as the baseline. **g**, Average reported number of contacts per 3 days in the 2-7 days after their test date. OR = odds ratio, LB = lower bound, UB = upper bound, CI = confidence interval, T = tested, P = positive, U = untested. **h**, Logistic regression analysis of factors contributing to users going to work in the 2-7 days after their COVID-19 test.

This overlap suggests that commonly used symptoms may not be sufficient criteria for evaluating COVID-19 infection. It has previously been reported that many people infected with SARS-CoV-2 are asymptomatic, mildly symptomatic, or in the presymptomatic phase of their presentation^21–23^ and therefore unaware that they are infected. In our dataset, on the day of their test, most users (73%) that tested PCR positive for SARS-CoV-2 reported feeling unwell with the common symptoms listed by the CDC (dry cough, shortness of breath, chills/shaking, fever, muscle/joint pain, sore throat, loss of taste/smell). However, 11.5% of positive users reported feeling unwell and exclusively reported symptoms not listed as common for COVID-19 by the CDC on the day of their test and, and 15.4% reported feeling no symptoms at all (Fig. 2g). Because of the commonly used symptom and occupation based screening criteria for receiving a PCR test and under-testing, this total of 36.9% likely underestimates the true fraction of asymptomatic, presymptomatic, and mildly symptomatic cases, which in Wuhan, China was estimated to be ∼80%^13^. A large number of asymptomatic cases were also observed in serological studies^24,25^. 48.9% of users testing negative for SARS-CoV-2 reported feeling unwell with most common COVID-19 symptoms, compared to an expected false negative rate of 20-30% for PCR-based tests of symptomatic patients^26^, again suggesting symptom presentation overlap with other diseases (Fig. 2g).

We investigated the symptoms that were most predictive of COVID-19 by exploring the distribution and dynamics of symptoms in PCR test (+) and (–) users around the test date. PCR test (+) users reported higher rate of common COVID-19 symptoms, including dry cough, fever, loss of appetite, and loss of taste and/or smell, than PCR test (–) users (Fig. 2h). Many PCR-tested users longitudinally reported symptoms in the app in an interval extending ±2 weeks from their test date (Extended Data Fig. 5). We used these data to examine the time course of symptoms among those who tested positive. In the days preceding a test, dry cough, muscle pain, and nasal congestion were among the most commonly reported symptoms. Reported symptoms peaked in the week following a test and declined thereafter (Fig. 2i). Taking the ratio of the symptom rates at each point in time between PCR test(+) and (–) users showed that the most distinguishing feature in users who tested positive was loss of taste and/or smell, as has been previously reported^10^ (Fig. 2j).

We next investigated medical and demographic factors associated with testing PCR positive for acute SARS-CoV2 infection, focusing on 3,829 users who took the V3 survey within ±2 weeks of their reported PCR test date (315 positive, 3,514 negative) (Fig. 3a, Extended Data Tables 2–6). These users are a subset of all the users who reported taking a test in the V3 survey, as some reported test results were outside this time window. To correct for selection bias of receiving a PCR test when studying the risk factors of a positive test result, we incorporated probability of receiving PCR tests as inverse probability weights (IPW) into our logistic model of PCR test result status (+/–) (Methods)^27^. As with the analysis of who received a test, the reported symptoms, loss of taste and/or smell was most strongly associated with a positive test result (OR: 33.17, 95% CI: [17.3, 67.94]). Other symptoms associated with testing positive included fever (OR: 6.27, 95% CI: [2.82, 13.70]) and cough (OR: 4.45, 95% CI: [2.83, 6.99]). Women were less likely to test positive than men (OR: 0.55, 95% CI: [0.38, 0.80]), and both Hispanic/Latinx users (OR: 2.59, 95% CI: [1.67, 3.97]) and African-American/black users (OR: 2.35, 95% CI: [1.25, 4.18]) were more likely to test positive than white users, highlighting potential racial disparities involved with COVID-19 infection risk. The odds of testing positive were also higher for those in high density neighborhoods (OR: 1.85, 95% CI: [1.15, 3.07]). Healthcare workers (OR: 1.92, 95% CI: [1.36, 2.73]) and other essential workers (OR: 1.69, 95% CI: [1.13, 2.52]) also had higher odds of testing positive compared to non-essential workers. Pregnant women were substantially more likely to test positive (OR: 6.30, 95% CI: [2.45, 14.68]). We note that this is a preliminary result based on a small sample of 48 pregnant women included in this analysis (9 test-positive, 39 test-negative) and will be reassessed as the app accumulates more users. Performing this analysis with and without correction for selection bias produced similar results (Fig. 3a).

Motivated by previous studies that reported high cluster transmissions occurred in families in China and Japan^28,29^, we explored household and community exposures as risk factors for users testing PCR positive. The odds of testing positive were much higher for those who reported within-household exposure to someone with confirmed COVID-19 than for those who reported no exposure at all (Methods) (OR: 19.10, 95% CI: [12.30, 30.51]) (Fig. 3a, Extended Data Table 5). This is stronger than comparing the odds of positive among those who reported exposure outside their household versus no exposure at all (OR: 3.61, 95% CI: [2.54, 5.18]). Further, the odds of testing PCR positive are much higher for those exposed in the household versus exposed outside their household or not exposed at all, after adjusting for similar factors (OR: 10.3, 95% CI: [6.7, 15.8]) (Extended Data Table 7). These results are consistent with previous findings that indicate a very high relative risk associated with within-household infection^29–33^. This is compatible with finding that other closed areas with high levels of congregation and close proximity, such as churches^34^, food processing plants^35^, and nursing homes^36^, have shown similarly high risk of transmission.

Developing models to predict who is likely to be SARS-CoV-2(+) from self-reported data has been proposed as a means to help overcome testing limitations and identify disease hotspots^10,11^. We used data from the 3,829 users who used the app within ±2 weeks of their reported PCR test results to develop a set of prediction models that were able to distinguish positive and negative results with a high degree of predictive accuracy on cross-validated data (Fig. 3b). We used the machine learning method XGBoost, which outperformed other classification methods (Extended Data Fig. 6). We considered: (1) a symptoms-only model, which included only the most common COVID-19 symptoms listed by the CDC; (2) an expanded model, which further incorporated other features observed in the survey; and (3) a minimal-features model, which retained only the four most predictive features (loss of taste and/or smell, exposure to someone with COVID-19, exposure in the household to someone with confirmed COVID-19, and exposure to household members with COVID-19 symptoms) (Methods, Extended Data Tables 8–11). The symptoms-only model achieved a cross validated AUC (area under the ROC curve) of 0.76 using data before and after a test, and AUC 0.69 using just the pre-test data. Expanding the set of features to include other survey questions substantially improved performance (cross-validated AUC 0.92 all data, 0.79 pre-test). In the minimal-features model, we were able to retain high accuracy (cross-validated AUC 0.87 all data, AUC 0.80 pre-test) despite only including 4 questions, one of which was a symptom and three referring to potential contact with known infected individuals. Restricting the observed inputs to the 1,613 users (89 positive, 1,524 negative) who answered the survey in the 14 days prior to being tested limited the sample size and reduced the overall accuracy, but the relative performance of the models was similar (Fig. 3b).

The fact that a fraction of SARS-CoV-2(+) users report no symptoms or only less common symptoms (Fig. 2g) raises the possibility that many infected users might behave in ways that could spread disease, such as leaving home while unaware that they are infectious. In spite of widespread shelter-in-place orders during the sample period, we found extensive heterogeneity across the US in the fraction of users reporting leaving home each day, with 61% of the responses from April 24 – May 12 indicating the user had left home that day (Fig. 4a). The majority (77%) of these users reported leaving for non-work reasons, including exercising; 19% left for work (Fig. 4b). Of people who left home, a majority but not all users reported social distancing and using face protection (Fig. 4c). This incomplete shutdown, and lack of total social and physical protective measures, coupled with insufficient isolation of infected cases, may contribute to continued disease spread.

Given the large number of people leaving home each day, it is important to understand the behavior of people who are potentially infectious and therefore likely to spread SARS-CoV-2. To this end, we further analyzed the behavior of people both reporting to be PCR test (+) or (–). There was an abrupt large increase in users reporting staying home after receiving a positive test result (Fig. 4d,e). Many, but not all, PCR test(+) users reported staying home in the 2-7 days after their test date (7% still went to work), whereas 23% of untested and 26% of PCR test(–) left for work (Fig. 4d,e). Similarly, 3% of PCR test(+) users reported going to work without a mask, in contrast with untested (12.7%) and PCR test(–) (10%) users (Fig. 4f). Positive individuals reported coming into close contact with a median of 1 individual over 3 days in contrast to individuals who tested negative or were untested, who typically came in close contact with a median of 4 people within 3 days (Fig. 4g). Regression analysis suggested that healthcare workers (OR: 9.6, 95% CI: [7.6, 12.1]) and other essential workers (OR: 7.0, 95% CI: [5.4, 9.1]) were much more likely to go to work after taking a positive or negative test, and PCR positive users were more likely to stay home (OR: 0.1, 95% CI: [0.1, 0.2]) (Fig. 4h, Extended Data Table 12).

We find evidence among our users for several factors that could contribute to continued COVID-19 spread despite widespread implementation of public health measures. These include a substantial fraction of users leaving their homes on a daily basis across the US, users who claim to not socially isolate or return to work after receiving a PCR test(+), self-reports of asymptomatic, mildly symptomatic, or presymptomatic presentation, and a much higher risk of infection for people with within-household exposure. That said, we note several limitations of this study. The HWF user base is inherently a non-random sample of voluntary users of a smartphone app, and hence our results may not fully generalize to the broader US population. Our results are based on self-reported survey data, hence may suffer from misclassification bias—particularly those based on self-reported behaviors. Moreover, a relatively small percentage of the US population has been prioritized for PCR testing, so any analysis of test results (or tested users) may be subject to selection bias. While we have attempted to correct for these biases via the inverse probability weighting approach (Methods), some residual bias may persist if there remain some unobserved factors related to underlying disease status and receiving a test.

Although there is enormous economic pressure on states, businesses, and individuals to be able to return to work as quickly as possible, our findings highlight the ongoing importance of social distancing, mask wearing, large-scale testing of both symptomatic and asymptomatic people, and potentially even more rigorous ‘test-trace-isolate’ approaches^37–40^ as implemented in Massachusetts, New York, and New Jersey to bend the infection curve^37–40^. Applying predictive models on a population scale will be vitally important to provide an “early warning” system for timely detection of a second wave of infections in the US and for guiding an effective public policy response. At an individual level, our data show that adding information beyond symptoms, such as household and community exposure, occupation, and demographics, is vital for identifying infected individuals from self-reported data. As testing resources are expected to continue to be limited, HWF results could be used to identify which groups should be prioritized, or potentially to triage individuals for molecular testing based on predicted risk. HWF’s integration of behavioral, symptom, exposure, and demographic data provides a powerful platform to address emerging problems in controlling infection chains, rapidly assist public health officials and governments with developing evidence-based guidelines in real time, and stop the spread of COVID-19.

## Data Availability

This work used data from the How We Feel project (http://www.howwefeel.org/). The software used in analysis will be made available on Github upon final publication of the paper. Researchers with IRB approval to perform research involving human subjects can apply to obtain access to data used in the analysis.

## Additional Material

## Acknowledgements

The How We Feel Project would like to thank operational volunteers Ari Simon, Ricki Seidman, Arun Ranganathan, Celie O’Neil-Hart, Debbie Adler, Divya Silbermann, Jack Chou, Lother Determann, Mark Terry, Rhiannon Macrae, Robert Barretto, Ron Conway, Sid Shenai, Tony Falzone, and Yurie Shimabukuro. We would also like to thank Andrew Tang for graphic design support. We are grateful to the HWF participants who took our survey and allowed us to share our analysis.

## Author contributions

W.E.A., H.A.-T., J.B., X.J., G.M., A.S., R.R., N.N., M.K. contributed to analysis. W.E.A., H.A.-T., J.B., X.J., and G.M., performed the majority of data cleaning, analysis, figures production, and wrote the manuscript with F.Z. and X.L. A.S. and R.R. performed household transmission and symptom type analysis. W.E.A. coordinated the analysis effort. A.P., C.D., A.K., J.I., T.H., E.C., C.L., M.C., H.B., W.L., R.M., R.P, B.S. designed and implemented the How We Feel application. B.C., M.T., J.O., C.S.G., O.S., G.K., and D.R.C. designed the survey, and provided feedback on app design and analysis. B.S. and F.Z. initiated the project. F.Z. and X.L. supervised all aspects of the work.

